# Comparing the efficacy of anti□infectious drugs for the treatment of mild to severe COVID-19 patients: a protocol for a systematic review and network meta-analysis of randomized clinical trials

**DOI:** 10.1101/2021.03.19.21253957

**Authors:** Dejene Tolossa Debela, Kidist Digamo Heraro

**Author notes:** Author of correspondence is to Dejene Tolossa Debela (DD): Center for Innovative Drug Development and Therapeutic Trials for Africa (CDT-Africa), College of Health Sciences, Addis Ababa University, P.O. Box 9086, Addis Ababa, Ethiopia, and.

## Abstract

**Background:** COVID-19 is a viral infection spreading at a great speed and has quickly caused an extensive burden to individuals, families, countries, and the world. No intervention has yet been proven highly effective for the treatment of COVID-19. Different drugs were being evaluated and reported through randomized clinical trials, and more are currently under trial. This review aimed to compare the efficacy of anti-infectious drugs with a comparator of the standard of care or placebo in patients with COVID-19.

**Methods and analysis:** Two independent review authors will extract data and assess a risk of bias using RoB2. Randomized controlled trials (RCT) that evaluate single and/or combined antiviral drugs recommended by WHO latest guideline for the treatment of COVID-19 will be included. We will search for Pub Med, the Cochrane Center for Clinical Trial database (CENTRAL), clinicaltrials.gov, etc. databases for articles published in the English language between December 2019 to April 2021. We will follow the Preferred Reporting Items for Systematic Review and Meta-Analysis Protocols (PRISMA-P) involving Network Meta-analysis guidelines for the design and reporting of the results. The primary endpoints will be time to clinical recovery and time to RNA negativity. The certainty of evidence will be evaluated using the GRADE extension of NMA. Data analysis will be performed using the frequentist NMA approach with netmeta package implemented in R.

**Ethics and dissemination:** There are no ethical considerations associated with this study as we will use publicly available data from previously published studies. We plan to publish results in open access peer-reviewed journals.

**PROSPERO registration number:** ID=CRD42021230919.

**Strengths and limitations of this study:**

- This will be the first systematic review and network meta-analysis to assess the efficacy specific to anti-infectious drugs category for for mild to severe patients of COVID-19.
- Its compliance with the Preferred Reporting Items for Systematic Review and Meta-Analysis for Protocols (PRISMA-P) involving network meta-analysis(NMA) will ensure the quality of reporting.
- Doing both pairwise meta-analysis and network meta-analysis (NMA) can comprehensively analyse direct and indirect comparison results of different anti-infectious drugs for COVID 19 will give more reliable conclusions aswell as the rank of those drugs.
- There is risk of heterogeneity and inconsistency, given the different anti-infectious drugs that will be included; however, we try to control intransitivity by carefully identifying the eligibility criteria depending on PICOS strategy and assess inconsistency using local as well as global approaches.
- The limitation of this study is it will not explore the economic benefits of these drugs.

## INTRODUCTION

In China, in December 2019, a novel coronavirus named severe acute respiratory syndrome coronavirus 2 (SARS CoV-2) caused an international outbreak of a respiratory illness called coronavirus disease 2019 [COVID-19](1). Since then, SARS-CoV-2 has spread globally, and COVID-19 has now been labeled a pandemic of international concern by the World Health Organization(2).

Coronaviruses are enveloped, positive-sense, single-stranded RNA virus genomes. The coronavirus encodes a nonstructural replicase polyprotein and structural proteins, including spike (S), envelope (E), membrane (M), and nucleocapsid (N)(3-5). The S protein on the surface of SARS-CoV is the most common target for the development of vaccines and therapeutics(6).

There are about thirty types of coronaviruses infecting mammals, birds, and other animals. Only seven of them infect humans(4, 7). Four of them usually cause mild diseases such as the common cold (HKU1; OC43; 229E; and NL63), whereas MERS-CoV, SARS-CoV, and now SARS-CoV-2 are likely to cause more serious diseases(5, 7). The main transmission way of SARS-CoV is from human to human by respiratory droplets(8-10).

COVID-19 disease clinical presentation can be from subclinical infection with mild (self-limiting respiratory tract illness) to severe (progressive pneumonia, multiorgan failure, and death) (11-14). Massive alveolar damage and progressive respiratory failure are the cause of death in severe covid-19 disease (12). Patients having comorbidities like people with chronic lung disease, serious heart disease, chronic kidney disease, elderly (above 65 years), and immunocompromised people are suspected to have the severe disease(15).

As of November 04, 2020, there were **47**,**362**,**304** confirmed patients, **1**,**211**,**986** confirmed deaths, and 219 countries, areas, or territories with COVID-19 according to the World Health Organization(16).

The only anti-viral drug FDA approved is Remdesivir yet for the treatment of COVID-19 in hospitalized patients (aged ≥12 years and weighing ≥40 kg)(17). To control the growing COVID-19 pandemic, we rely on quarantine, isolation, and infection-control measures preventing the spread of disease as well as oxygen and mechanical ventilation as supportive care for infected patients(18). Currently, there are many drugs exist that are being under assessment for patients with COVID-19: example, remdesivir (used to treat Ebola virus disease and Marburg virus infections), lopinavir and ritonavir (used to treat HIV/AIDS), chloroquine phosphate or hydroxychloroquine (used to treat malaria), tocilizumab (used to treat rheumatoid arthritis), corticosteroids, stem cells, and other types of interventions(19).

Several randomized clinical trials are underway. According to an online global COVID-19 clinical trial tracker available at www.covid19-trials.org, there are currently 2462 trials registered worldwide as about 20% of them are in the US.

Although the mortality rate is concerning, the high transmissibility of the disease is much more alarming. Even if a low percentage of patients need hospitalization, the rapid spread of the disease and a large number of people infected has overwhelmed the healthcare systems worldwide. To decrease the spread, severe social distancing measures, travel restrictions, closures of schools, and many businesses are taking an unprecedented socioeconomic and psychological toll. Therefore, COVID-19 has caused an enormous impact on people’s quality of life and posed far-reaching threats, especially to the economy, health, and the sustainability of healthcare systems(18).

There have been many efforts done to identify effective drug treatment for covid-19 but, evidence for effective treatment remains limited. It is, therefore, an urgent need of investigating the most effective drugs to slow the progression of the disease and unburden the health care systems. Although extraordinary efforts have been made on research regarding pharmacological interventions, none have proven most effective. Therefore, this systematic review and network meta-analysis aim to synthesise existing evidence to compare the efficacy and safety as well as identify the best drug among different antivi-infectious drugs category for the treatment of mild to severe patients with COVID-19.

## METHODS

This research is designed and will be reported by a systematic review involving a network meta-analysis that will comply with the Preferred Reporting Items for Systematic Review and Meta-Analysis Protocols (PRISMA-P) 2015 guidelines(20). This protocol has been registered at the PROSPERO 2021 database,ID=CRD42021230919.

### Data sources and searches

We will search databases from December 2019 to April 2021. We will conduct search for PubMed, the Cochrane Center for Clinical Trial database (CENTRAL), clinicaltrials.gov, clinicaltrialsregister.eu, chictr.org.cn, covid-19.cochrane.org, and covid-evidence.org. databases for articles published worldwide in the English language. The search will be done according to guidance provided in the Cochrane Handbook for Systematic Reviews of Interventions. The search will be limited to human studies, published in English languages until April 2021. We will check for any unidentified randomized clinical trials from reference lists of relevant trial publications. The authors of the included trials will be contacted by email asking for unpublished randomized clinical trials. We will do a thorough search strategy using the Mesh terms and key words including SARS-CoV-2, COVID-19, COVID-19 serotherapy, SARS-CoV-2 variants, Antiviral Agents, Hydroxychloroquine, Remdesivir, GS-441524, Lopinavir, Lopinavir-ritonavir drug combination, favipiravir, Ivermectin, chloroquine, azithromycin, Randomized Controlled Trial and RCT. Table 1 summarises the search strategy that we will used in PubMed database (Table 1).

**Table 1.** Search strategy for the PubMed database.

| SN | Search terms | Total articles |
| --- | --- | --- |
| #1 | "Antiviral Agents"[Mesh] OR "Antiviral Agents" [Pharmacological Action] OR "Hydroxychloroquine"[Mesh] OR "remdesivir" [Supplementary Concept] OR "GS-441524" [Supplementary Concept] OR "Lopinavir"[Mesh] OR "lopinavir-ritonavir drug combination" [Supplementary Concept] OR "favipiravir" [Supplementary Concept] OR "Ivermectin"[Mesh] |  |
| #2 | "SARS-CoV-2"[Mesh] OR "COVID-19"[Mesh] OR "COVID-19 serotherapy" [Supplementary Concept] OR "SARS-CoV-2 variants" [Supplementary Concept] |  |
| #3 | "Randomized Controlled Trial" [Publication Type] OR "Randomized Controlled Trials as Topic"[Mesh] |  |
| #4 | #1 AND #2 AND #3 |  |

### Eligibility criteria

We will identify eligible studies through the PICOS(participants, interventions, comparison, outcomes, and study designs) format(21)

#### Types of participants

We will include all patients diagnosed with laboratory-confirmed, mild to severe COVID 19 patients of both sexes and all ages with any comorbidities.

#### Types of interventions

We will include any anti-infectious drugs used to treat COVID-19 recommended by WHO’s latest guideline including Hydroxychloroquine, Remdesivir, Lopinavir, Lopinavir-ritonavir, favipiravir, Ivermectin, chloroquine, azithromycin etc.

#### Types of comparators

We will include the standard of care, placebo, or another drug group.

#### Types of outcome measures

#### Primary outcome

- Time to clinical improvement and time to viral clearance(RNA negativity)

#### Secondary outcomes

- Mortality rate, length of hospital stay, rate of patients need for oxygen therapy and adverse events (AE).

#### Types of studies

We will include only randomized clinical trials (RCT) that compared efficacy of anti-infectious drugs for treatment of COVID 19 against standard care or placebo or other medication.

### Selection of studies

Endnote software version X7 will be used to import the research articles from the electronic databases and duplicates will be removed. Two review authors will independently screen titles and abstracts based on pre-specified eligibility criteria and retrieve all relevant full-text study reports. Any disagreements between two review authors will be resolved through discussion, or if required, they will consult a third person. The screening and selection process will be reported in a PRISMA flow chart as summarized by Figure 1.

**Figure 1:**
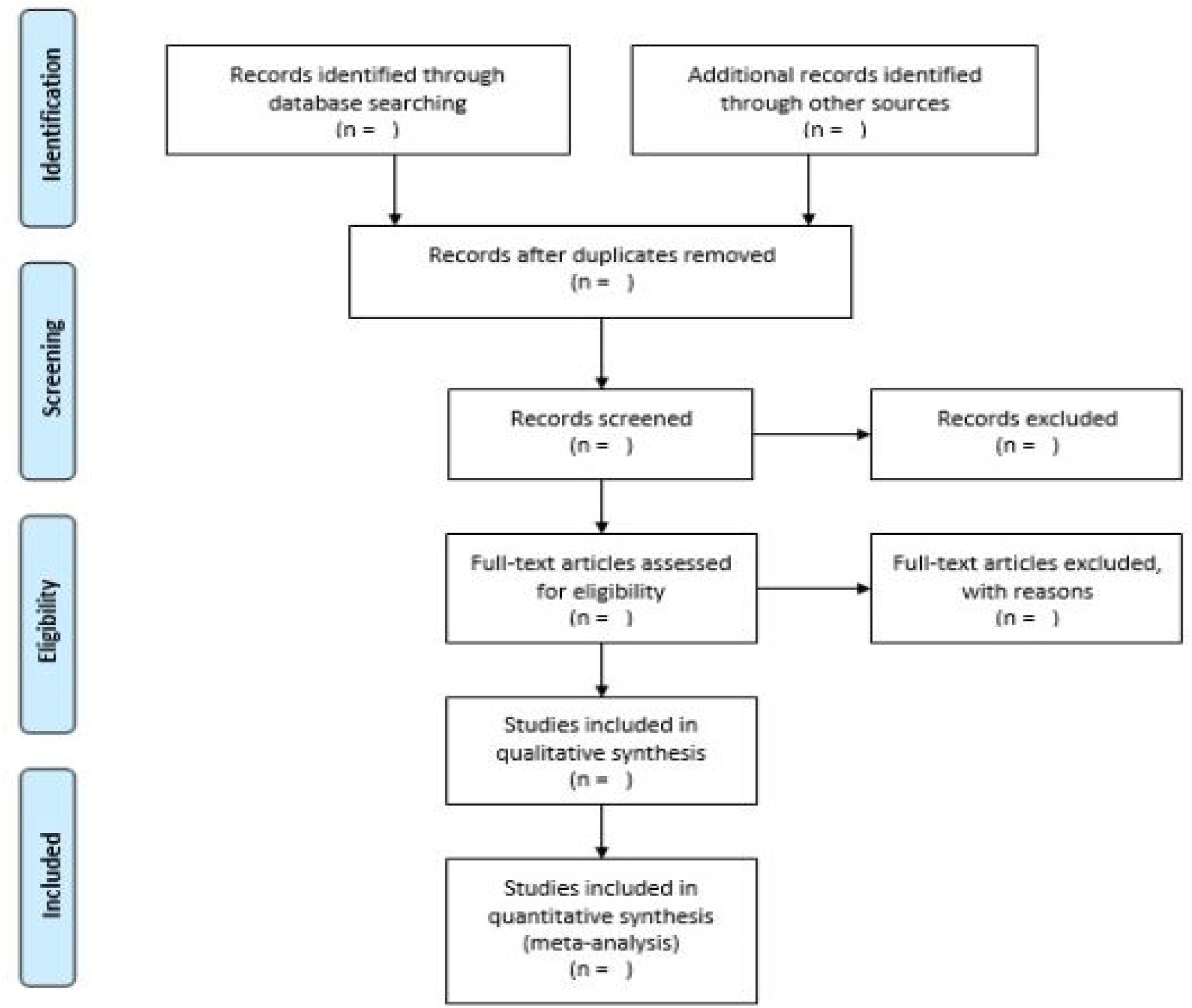
PRISMA flow chart.

### Data extraction

The data will be extracted independently by 2 reviewers using a predefined format. Disagreements will be resolved by discussion. The two review authors will evaluate all available data simultaneously to maximize data extraction by assessing duplicated publications and companion papers of a trial together. We will name each trial after the first author and year of the publication. The information will be collected include participants (demographic and clinical characteristics), the pharmacological treatment (name of the drug, treatment duration, dose), time points used for the assessments, number of patients lost to follow-up (in each group), reasons for loss to follow-up, missing data (intention-to-treat or per protocol), sources of funding, possibility of a conflict of interests, adverse events, outcome measures (primary and secondary outcomes), protocol deviations. Also, Relevant information such as title, author name, year of publication, publication status, study design, study setting, follow-up period, sample size, funding of the trial or sources of support, baseline characteristics of study subjects, will be extracted. The trial authors will be contacted by email to specify any missing data, which may not be reported sufficiently or not at all in the publication.

### Assessment of risk of bias

The risk of bias assessment will be based on the Cochrane Risk of Bias tool version 2 (Rob 2) as recommended in The Cochrane Handbook of Systematic Reviews of Interventions(22). We will evaluate the methodology to reduce the risk of bias across the following five domains (Bias from the randomization process, deviation from intended interventions, missing outcome data, measurement of outcomes and selective reporting of results). The risk of bias of each trial will be judged by two independent authors as low risk, some concerns, and a high risk of bias. The disagreements will be resolved by discussion between the two authors.

### Statistical analysis

We planned to do all statistical analyses using R version 4.0.3 software for Window 10. Odds ratios (OR) with a 95% confidence interval (CI) for dichotomous outcomes and Mean differences (MDs) with 95% CI for continuous outcomes data will be used to estimate the relative treatment effects of the competing interventions. We will convert other forms of data into MDs using standard conversion formula. For outcome variables reported in different scales, we will use standard mean differences with 95% CIs. Other binary outcome data will be converted into OR. Estimation of the ranking probabilities will be done for all included anti-infectious drugs of being at each possible rank for each intervention. Surface under the cumulative ranking curve (SUCRA) and mean ranks will be used to obtain a treatment hierarchy.

### Meta analysis and network meta analysis

We will undertake the meta-analyses according to the Cochrane Handbook of Systematic Reviews of Interventions(23) using R version 4.0.3 software for every treatment comparison with at least two studies. Effect sizes of individual studies and any pooled estimates of effect will be presented in tables and graphically as forest plots.We will use forest plots to visually evaluate any sign of heterogeneity. Then we assess the presence of statistical heterogeneity using I2 statistic. Substantial heterogeneity (I^2^> 50%)(24-26). We planned to perform network meta-analysis using a frequentist NMA approach and a random effects model for each treatment comparison, using the netmeta package version 1.2-1 implemented in R version 4.0.3 software for Window 10, if the assumption of transitivity is fulfilled. We will categorize the network nodes as follows: 1. **Chloroquine, 2. Hydroxychloroquine** 3. **Lopinavir/Ritonavir**, 4. **Remdesivir**, 5. **Ivermectin 6. Favipiravir, 7. The combinations** of those drugs. We will assess inconsistency globally across the whole network. If evidence of inconsistency is found, we will assess locally (a node splitting approach) to identify possible areas of local inconsistency and, if sufficient data exist, run network meta-regression. We will extend the analysis to all closed loops assuming a loop-specific heterogeneity and examine the estimates of inconsistency together with 95% confidence intervals for each loop using a graphical representation. Any orderings of treatment hierarchy will be estimated in primary outcomes and present treatment rankings with P-score using Netrank of the R package netmeta.

We will conduct a sensitivity analysis for all studies assessed as being low risk of bias and high risk of bias to test the robustness of our data. We will do subgroup analysis using network meta regression in the primary outcomes influenced by available variables: age, sex, comorbidities, and disease severity (mild, moderate, and severe).

Confidence in the evidence will be assessed using the GRADE working group recommendations and the CINeMA software and classify evidence as *high, moderate, low, or very low* certainty(27, 28). This rating method follows steps required are from 1) direct, 2) indirect, 3) NMA evidence, and 4) direct and indirect comparisons. Assessments of the evidence will be presented using the six domains: (study limitations, indirectness, inconsistency, imprecision, publication bias, incoherence)(28). The results will be presented in a ‘NMA SoF table.

### Dealing with missing data

We will contact all trial authors to obtain clarification for any relevant missing data. If the authors will not respond imputation method will be used.

### Reporting bias

We will use a the comparison-adjusted and contour-enhanced funnel plot and Egger’s test to visually assess publication bias if ten or more trials will be included(24). We also use the adjusted rank correlation(29, 30).

### Patient and public involvement

There was no patient or public involvement in this systematic review and network meta-analysis.

### Ethics and dissemination

There are no ethical considerations associated with this study as we will use publicly available data from previously published studies. We planned to publish results in open access peer reviewed journals.

### Amendments

The protocol for this study will be amended as necessary.

## Data Availability

data is available online

## Acknowledgements

The authors would like to acknowledge the Center for Innovative Drug Development and Therapeutic Trials for Africa (CDT-Africa), College of Health Sciences, Addis Ababa University which funds this study.

## Author contributions

DTD conceived the content, wrote the paper, and approved the final version. HKD conducted the preliminary search, copyedited and revised the manuscript. Both authors have read and approved the manuscript.

## Patient consent for publication

Not required

## Provenance and peer review

Not commissioned; externally peer reviewed.

